# Longitudinal prediction of motor dysfunction after stroke: a disconnectome study

**DOI:** 10.1101/2021.12.01.21267129

**Authors:** Lilit Dulyan, Lia Talozzi, Valentina Pacella, Maurizio Corbetta, Stephanie J Forkel, Michel Thiebaut de Schotten

**Author notes:** contributed equally (Stephanie J Forkel, Michel Thiebaut de Schotten).

## Abstract

Motricity is the most commonly affected ability after a stroke. While many clinical studies attempt to predict motor symptoms at different chronic time points after a stroke, longitudinal acute-to-chronic studies remain scarce. Taking advantage of recent advances in mapping brain disconnections, we predict motor outcomes in 62 patients assessed longitudinally two weeks, three months, and one year after their stroke. Results indicate that brain disconnection patterns accurately predict motor impairments. However, disconnection patterns leading to impairment differ between the three time points and between left and right motor impairments. These results were cross-validated using resampling techniques. In sum, we demonstrated that while some neuroplasticity mechanisms exist changing the structure-function relationship, disconnection patterns prevail when predicting motor impairment at different time points after stroke.

## Introduction

Stroke is one of the most significant disorders worldwide both in terms of mortality and long-term disabilities (Feigin et al. 2021). Despite considerable research into the risks and treatments of cerebrovascular diseases (Gurol and Kim 2018), every 3 seconds, someone suffers a stroke, resulting in various cognitive and motor dysfunctions (www.world-stroke.org). Motricity is understood as a motor impulse sent efferently down a nerve toward a muscle. This motor function is the most commonly affected ability after stroke (∼88%, Aqueveque et al. 2017). Motor deficits post-stroke occur secondary to a vascular rupture (haemorrhage) or an occlusion (ischemia) leading to decreased blood flow in one of the arteries that supply the motor network in the brain. This network includes the primary motor and somatosensory cortices, premotor regions in frontal and parietal cortex, and the basal ganglia. The main outflow tract is the corticospinal tract (CST, Bhuiyan et al. 2014) that descends from the cortex through the internal capsule, the cerebral peduncle, the brainstem down into the spinal cord. Post-stroke motor deficits such as partial paralysis or muscular weakness (hemiparesis) of the upper (e.g. hands, arms) or lower extremities (e.g. legs, foot) are present in about 88% of stroke patients at the acute stage. Longitudinally, merely 12% of patients demonstrate a full functional recovery after rehabilitation (Aqueveque et al. 2017). Although several rehabilitation programmes have been developed (e.g., The Queen Square Upper Limb (QSUL) Neurorehabilitation programme, Kelly et al. 2020), at present, there is no reliable prediction of recovery. This absence of reliable predictions can lead to frustration and reduced motivation in patients and family members, which is unfavourable to rehabilitation success (Kusec et al. 2019). Since proper and early interventions are crucial for a successful recovery (Coleman et al. 2017), early recovery prediction will likely help clinicians adjust their rehabilitation efforts to significantly improving motor functioning after a stroke. Hence, accurate predictions may result in more efficient and successful recovery of patients long-term.

Using various types of biomarkers, a wealth of clinical studies attempted to predict motor symptoms at the acute (first seven days), subacute (within one week to three months), and early and late chronic phases (starting at three months after onset) after stroke (Boyd et al. 2017; Connell et al. 2018; Stinear 2017; Lin et al. 2018). This body of work showed that the quality of motor abilities after stroke is associated with i) the severity in the clinical behavioural baseline scores assessed at the acute stage (Nijland et al. 2010), ii) the presence and amplitude of motor evoked potentials triggered with transcranial magnetic stimulation (TMS, Feys et al. 2000), iii) the volume and location of the lesion calculated with structural magnetic resonance imaging (Schiemanck et al. 2006), iv) the structural integrity of the cortico-spinal tract detected with diffusion-weighted imaging (DWI, Puig et al. 2011; Stinear 2017), task-related cortical activity and resting-state functional connectivity measured with functional MRI (fMRI, Rehme et al. 2011; Thiel and Vahdat 2015; Carter et al. 2010; Carter et al. 2012). Although most of these studies demonstrate a strong statistically significant relationship between behavioural motor scores and distinct biomarkers that can explain up to 96% variance of the data (Granziera et al. 2012), they do not validate the results of their models in an independent sample of patients which reduces the ecological validity and generalisability of these results. A systematic review (Kim and Winstein 2017) showed that only 8 out of 71 studies aiming to predict motor outcomes performed a resampling technique (e.g. k-fold cross-validation) to evaluate their models. These methods fight against a common problem in machine learning known as overfitting. A model perfectly describes the known data - training set, but it performs poorly on new observations - testing set (Ying 2019). Those algorithms that demonstrate the perfect ability to capture the variability in data without applying resampling methods may fail to generalise as a universal model to predict recovery (Heil et al., 2021). Thus, testing the model for out-of-sample predictions is crucial for translating the knowledge from research to a clinically useful tool to maximise patient benefits. As such, the absence of externally validated models makes their performances unreliable in predicting motor outcomes of new individual patients (Berrar, 2019) and further high-quality statistic studies are required in order to identify reliable brain markers for motor deficits after a stroke.

Many efforts have been dedicated to studying lesion localisation and associating it with motor disabilities (see review Schiemanck et al. 2006). It is undeniable that these symptoms can also occur with lesions to the grey or white matter (Catani and ffytche 2005; Carrera and Tononi 2014; Thiebaut de Schotten and Foulon 2018; Thiebaut de Schotten et al. 2008). For example, in spinal cord disconnections, the information processed in the intact motor cortex cannot be conducted to the relevant part of the body, causing motor paralysis (Tidoni et al. 2015). In the 19th century, this type of interruption of motor function distant from a lesioned region was discussed by a pioneering neurologist, Constantin von Monakow (1853-1930, von Monakow 1914), and referred to as *diaschisis cerebrospinalis* (Finger et al. 2004). However, a proper statistical measure of this theoretical concept was not possible thus far.

Advanced neuroimaging methods, including DWI that measures water displacement in brain tissue, opened a new avenue into exploring white matter connections (Moura et al. 2019; Catani and Thiebaut de Schotten 2008). Nowadays, the lesion load (the proportion of destruction) in the CST is one of the most popular predictors of motor disabilities and is interpreted as a measure of diaschisis (Carrera and Tononi 2014; Koch et al. 2021). However, lesion load in the CST does not account for the dysfunction of other distant brain regions. In other words, lesion load analysis is not an accurate operationalisation of diaschisis (Thiebaut de Schotten et al. 2014). Limiting our analysis to the lesion location and neglecting possible interruptions of functions in other brain structures might miss valuable information to understand behavioural outcomes after stroke. The disconnection analysis has been proposed recently as a possible solution to this problem (Foulon et al. 2018; Kuceyeski et al. 2013). Disconnection analyses consider a probability of dysfunctions in other brain areas. For example, if a patient has a lesion in a subcortical region (e.g. posterior limb of the internal capsule), the disconnection analysis will provide us with a probability of interruption of the white matter connection (e.g. CST) that links the subcortical (internal capsule) and cortical regions (e.g. primary motor cortex). When comparing those methods, the disconnection analysis often explains more variance in data than lesion load analysis (Corbetta et al. 2015; Hope et al. 2016). The disconnection analysis (Conrad et al, 2022; Hajhajate et al, 2022; Forkel et al. 2022); Souter et al, 2022; Silvestri et al, 2022; Sperber et al, 2022) is different from the lesion network symptom mapping methods that estimate from a normative dataset the functional connectivity impaired by a lesion (Boes et al. 2015; Bowren et al, 2022; Cototvio et al, 2022). Thus, prediction studies may benefit from the redirection of the focus in clinical research from lesion localisation analysis to the analysis of the disconnections where remote alterations of a focal damage to distant regions can also be considered as a contributor to a network of structures and functions.

Another issue in the literature that may downplay the full potential of predicting motor symptoms is the considerable variation in motor assessments. Each clinical test utilised in symptom prediction studies focuses only on one aspect of motor dysfunction (Kim and Winstein 2017), which is not capturing the whole complexity of motor disabilities (Cheung et al. 2012), making both the prediction and comparisons between studies challenging. Corbetta et al. (2015) partially overcame this limitation by applying a principal component analysis that explained a great majority of variability (77%) across all motor tasks and patients with just two components (i.e. left and right side of the body). Extracting common patterns of motor outcomes from different assessments and eliminating the redundancy in the data should increase the performance of predictive models in the future (Corbetta et al. 2015).

Recent work considered the main limitations outlined above and conducted a ridge regression model that penalizes weights of the predictors by applying second-order penalty terms (i.e. L2-normalisation) to avoid overfitting. This model explained up to 37% and 42% of the variability in motor outcomes at the 2 weeks for left motor recovery (i.e. right hemisphere lesion) and right motor recovery (i.e. left hemisphere lesion), respectively (Salvalaggio et al. 2020). While these results are encouraging, the predictions reported are limited to 2 weeks after the stroke and do not extend to the long-term prognosis of motor abilities.

This study aimed to predict acute-to-chronic motor outcomes in 62 stroke patients two weeks, three months, and one year after stroke from the lesion defined at the acuate stage (two weeks post stroke). Using a cross-validation longitudinal study design and a disconnection analysis should overcome the limitations of current predictive models in stroke. We modelled the pattern of brain disconnections based on the Disconnectome (Thiebaut de Schotten et al. 2020), which characterises brain disconnections along 46 anatomical dimensions. The predictions were calculated using ridge regression models with nested cross-validation resampling and yielded medium to large effect sizes.

## Methods

### Participants

Sixty-two patients (age: M = 53.7, SD = 10.7, range 22-83 years; 34 males; 57 right-handed, see Table 1) met the inclusion criteria: All patients were older than 18 years, presented with first-ever ischemic (90%) or haemorrhagic (∼10%; 6 patients) stroke and behavioural deficits as assessed by a neurological examination. Patients who had a history of neurological or psychiatric presentations (e.g. transient ischemic attack), multifocal or bilateral strokes, or had MRI contraindications (e.g. claustrophobia, ferromagnetic objects) were excluded from the analysis (n = 131 patients, see the enrollment flowchart in the supplementary materials from Corbetta et al. 2015). We further limited our analysis to the patients whose motor functions were systematically assessed at 2 weeks, 3 months, and 1 year after their stroke for optimal longitudinal comparisons. This study was approved by the Washington University in Saint Louis Institutional Review Board and all participants gave their signed informed consent.

**Table 1.** Demographics of patients

|  | **Statistic** | **Min** | **Max** |
| --- | --- | --- | --- |
| **Age (mean/SD)** | 53.73/ 10.66 | 22 | 83 |
| **Education (mean/SD)** | 13.66/ 2.76 | 5 | 20 |
| **Handedness (% right-handed)** | 91.94 |  |  |
| **Sex (% female)** | 45.16 |  |  |

Abbreviations: SD = standard deviation

### Motor abilities assessment

Motor functions were examined for the upper and lower extremities. For upper extremities, active range of motion against gravity was measured by goniometer at shoulder flexion and wrist extension (Dreeben 2008). During the examination of shoulder flexion, patients are asked to raise their arm against gravity as high as possible. The movement amplitude is recorded as the angle between the goniometer centred on the shoulder and the lateral torso. The wrist extension examination requires patients to sit with their arm on the table in a resting position with their palms down, and they are asked to bend back their wrist against gravity. Wrist extension is measured as the angle between the goniometer centred on the wrist and the forearm.

Grip strength was measured using a dynamometer (Demeurisse et al. 1980). Each patient’s examined arm was placed with the elbow flexed at 90°. Their fingers flexed for a maximal contraction over the dynamometer handle, while the forearm and wrist were kept in a neutral position. The patients were asked to take a breath while exerting the maximum grip effort during three consecutive trials. The strength score is recorded in kilograms, and the total score is calculated as the average over three trials (Fess and Moran 1981).

The patients’ dexterity and bimanual coordination was measured with the 9-Hole Peg Test (pegs/second) assessing gross movements of arms, hands, and fingers, and fine motor extremity. The test includes a one-piece board with a concave folded dish containing nine pegs next to a 9-holes matrix for the pegs. The task instructions require patients to place and remove the nine pegs one at a time and in random order as quickly as possible (Mathiowetz et al. 1985; Oxford Grice et al. 2003). The final score is calculated as the time in seconds elapsed from the touch of the first peg to when the last peg is placed back into the dish.

The Action Research Arm Test (ARAT, van der Lee et al. 2001) assesses the ability to perform purposeful movements with the upper limb extremities. Patients had to grasp, grip, pinch objects of different weights and shapes and perform gross motor movements. The *ARA*T’s four subtests have 19 items in total. Each item is rated on a four-point scale (0-3) where higher scores indicate better performance. If the patient scores <3 on the first item, the examiner advances to the second item two (the most accessible item). If the score for the second item is 0, the rest of the items will automatically be scored as 0 for and the test is stopped. If the patients score <3 on the first item but >0 on the second item, the remaining items are administered (Lyle 1981).

For lower extremities, a combined walking index (Kempen et al. 2011; Perry et al. 1995), left/right total motricity index, ankle dorsiflexion goniometry for left/right active range of motion against gravity was recorded (Dreeben 2008).

### MRI acquisition and preprocessing

#### 1. MRI scan acquisition

Neuroimaging was performed on a Siemens 3T Tim-Trio scanner at the School of Medicine of the Washington University in St. Louis. All structural scans were collected 2 weeks after the stroke and included (1) a sagittal MP-RAGE T1-weighted image (repetition time = 1950 msec, echo time = 2.26 msec, flip angle = 9 degrees, voxel size = 1.0 × 1.0 × 1.0 mm, slice thickness = 1.00 mm); (2) a transverse turbo spin-echo T2-weighted image (repetition time = 2500 msec, echo time = 435 msec, voxel-size = 1.0 × 1.0 × 1.0 mm, slice thickness = 1.00 mm); and (3) a sagittal FLAIR (fluid attenuated inversion recovery) (repetition time = 7500 msec, echo time = 326 msec, voxel-size = 1.5 × 1.5 × 1.5 mm, slice thickness = 1.50 mm).

#### 2. Lesion segmentation (native space)

Lesions were manually segmented on the T1-weighted MRI images using the Analyze biomedical imaging software system (Robb and Hanson, 1991). Two board-certified neurologists (Drs Corbetta and Carter) reviewed all segmentations blinded to the individual behavioural data.

#### 3. Spatial normalisation (MNI152)

To align T1-weighted MRI scans of patients to a standard stereotaxic space (Montreal Neurological Institute space, MNI152 Grabner et al. 2006), it is necessary to first address the issue of space deformation caused by brain lesions during spatial normalisation (Brett et al. 2001; Ripolles et al. 2012; Volle et al. 2013). An enantiomorphic approach was implemented in the current data analysis: the native-space lesions were replaced with healthy tissue of the same region in the contralateral hemisphere (Nachev et al. 2008). Subsequently, affine and diffeomorphic deformations were applied to co-register scans and lesions to the MNI152 space using the Advanced Normalization Tool (ANTS, Avants et al. 2011; Klein et al. 2009). These analyses are available as part of the ‘Normalisation’ part of BCBtoolkit (Foulon et al. 2018; http://toolkit.bcblab.com).

#### 4. Generation of disconnection maps

Methodological details are available from Thiebaut de Schotten et al. (2020). In brief, each lesion serves as the input for the BCBtoolkit’s Disconnectome tool that computes maps of white matter pathway disconnection probabilities and its impact on loss of function (Foulon et al. 2018; http://toolkit.bcblab.com).

Probabilities of white matter pathways were derived from a normative population of 163 healthy controls (44.8% males) using a diffusion-weighted imaging dataset acquired on a 7T scanner as part of the Human Connectome Project (Vu et al. 2015). The pattern of brain areas that were disconnected in each stroke was subsequently characterised by measuring the average level of disconnection in subcortical areas and areas derived from a multimodal atlas of the brain surface (Glasser et al. 2016). Subcortical areas included manually defined by MTS and included the thalamus, the putamen, the pallidum, the hippocampus, the caudate nucleus, and the amygdala. Hence, for each of the 62 patients in this study, the disconnection probability of 372 grey matter structures (186 structures in each hemisphere: 180 cortical and 6 subcortical) was obtained.

### Dimensionality reduction with Principal Component Analysis

#### 1. Behavioural components

To summarise behavioural measurements data while keeping as much variability as possible and minimising noise, behavioural components were extracted as described in Corbetta et al. (2015). In brief, an oblique rotation principal component analysis was applied to the motor scores obtained with different neuropsychological tests described above. Two components (left and right side of the body) resulted from the analysis explaining 77% of the observed variance in the data. The analysis was computed in Matlab (MathWorks Inc.).

#### 2. Disconnection map components

The Disconnectome consists of 46 components, where 30 components have already been shown to capture more than 90% of the variance in the distribution of disconnection maps in stroke (Thiebaut de Schotten et al. 2020). These components were derived from an independent normative dataset of 1333 disconnection maps of ischemic stroke patients (M = 63.89, SD = 15.91, range 18-97 years; 56.1% males) fully described in (Xu et al. 2018) and (Thiebaut de Schotten et al. 2020).

### Estimation of the component’s scores

We used the Disconnectome 46 components to describe the disconnection patterns in our sample of 62 patients. Linear regression with the Disconnectome components (i.e. 46 independent variables: predictors: matrix of 372 regions x 46 components) was performed in RStudio (v.1.3.1093; RStudio Team, 2020) to predict the disconnection pattern (i.e. the dependent variable: matrix of 372 regions x 1 patient) for each patient. In total, 62 linear regression were performed. In doing so, unstandardised beta coefficients for each predictor corresponded to the component’s scores of each patient (see Supplementary material Figure 1 for the workflow). Adjusted r-squared values represented the percentage of each patient’s disconnection pattern variance explained by the 46 components. The linear regression analysis demonstrated that the Disconnectome (46 principal components) was able to capture the variance in the disconnection pattern of each patient from the longitudinal dataset. The mean adjusted r-squared across the group is 0.94, with a standard deviation of 0.076 (https://github.com/lidulyan/Estimate_CompScore_lm)

**Figure 1.**
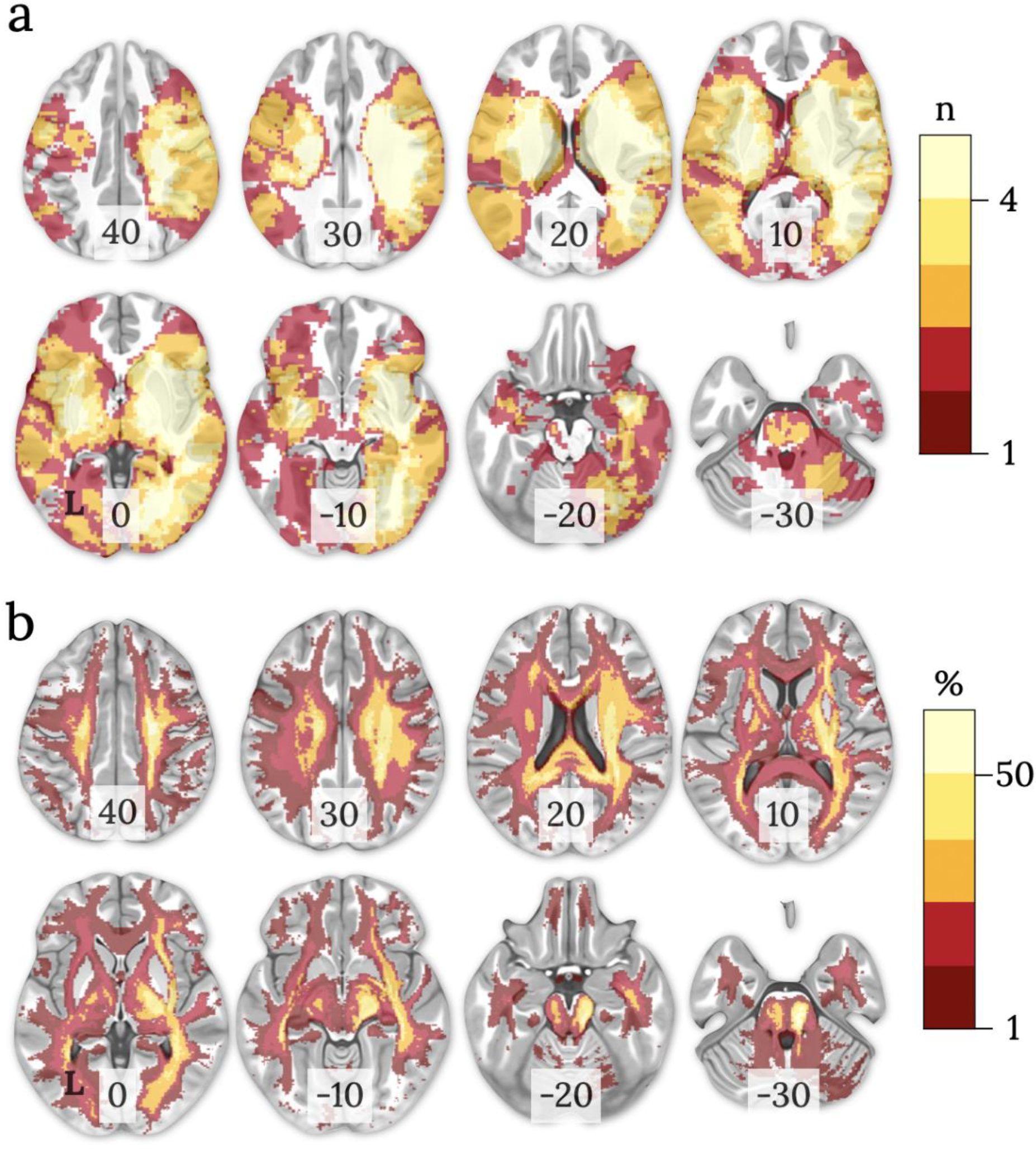
Distribution of (a) lesions and (b) disconnection maps in 62 stroke patients in the MNI152 space (z coordinate indicated on each slice, neurological view: left=left). The colour bars represent a) the number of overlapping lesions and b) the probability of disconnection. L: left hemisphere; n: number.

### Statistical analysis

The analyses for the anatomical prediction of the motor outcome were carried out in RStudio (v.1.3.1093; RStudio Team, 2020). Backward/forward hierarchical linear regressions, backward/forward stepwise regressions, ridge and lasso regressions were performed and the results between different approaches were compared. Each regression used the patients’ estimated scores of each component (i.e. beta coefficients from the linear regression) to predict two principal components of motor scores (the left and right side of the body, n = 2) at two weeks, three months, and one year after the stroke (n = 3 time points) resulting in 6 regression models per each type of regression.

This procedure is available as supplementary code with the manuscript (see https://github.com/lidulyan/Hierarchical-Linear-Regression-R- and https://github.com/lidulyan/Stepwise-Lasso-Ridge).

#### 1. Backward/Forward hierarchical linear regressions

In the backward hierarchical linear regressions (bHLR), predictors were eliminated one by one, starting from the last one. If the comparison between two consecutive models was statistically significant then the eliminated predictor was retained in the model. For example, a model with components 1-46 is compared with a model with components 1-45. If the comparison is not significant then we remove component 46 from the model. The goodness of fit of consecutive linear models was compared statistically using an F-test. The process was repeated until no significant difference could be identified between the two consecutive models. In the case of the forward hierarchical linear regressions (fHLR), the predictors were added one by one, starting from the first one. The last comparison is the optimised model vs. the optimised model without Component#1.

To avoid inflation of the significance of our results (i.e. overfitting), the hierarchical linear regression analyses were performed on 78% of the original dataset (training set), and the model accuracy on the testing set (22% from the original dataset was assessed with R.

#### 2. Backward/Forward stepwise regressions

In the backward stepwise regressions, the least significant predictors were iteratively removed from the model until the model contains only statistically significant predictors. In the case of the forward stepwise regressions, the most significant predictors were iteratively added to the model until the model contains only statistically significant predictors.

The allowed maximum number of predictors to be included in the model was varied from 1 to 46. The optimal hyperparameter value was selected on the training set (80% of data: 50 patients) based on the results of the leave-one (patient)-out cross-validation (LOOCV) procedure: (train on 59, test on 1). The model with the smallest prediction error (root-mean-square error (RMSE) value) was considered as the best model and it was tested on the unseen during the training procedure data (20% of data: 12 patients). Model accuracy on the testing set was assessed with R.

#### 3. Lasso regressions

Lasso regression is another type of linear model that performs variable selection with L1-regularisation which results in the shrinkage of the model where some coefficients become 0, and therefore eliminated from the model. The larger the penalty (L1-regularisation: λ1), the more coefficient values are 0.

The optimal hyperparameter value (λ1) was selected on the training set (80% of data: 50 patients) based on the results of the leave-one (patient)-out cross-validation (LOOCV) procedure: (train on 59, test on 1). The model with the smallest prediction error (root-mean-square error (RMSE) value) was considered the best model and it was tested on the unseen during the training procedure data (20% of data: 12 patients). Model accuracy on the testing set was assessed with R.

#### 2. Ridge regressions

Similar to Lasso, Ridge regression penalises the less important model coefficient values in order to prevent the model from overfitting the data, however, it never makes them 0.

L1-regularisation (λ2) is a hyperparameter in the ridge regression and it was tuned on the training set (80% of data: 50 patients) based on the results of the leave-one (patient)-out cross-validation (LOOCV) procedure: (train on 59, test on 1). The model with the smallest prediction error (root-mean-square error (RMSE) value) was considered the best model and it was tested on the unseen during the training procedure data (20% of data: 12 patients). Model accuracy on the testing set was assessed with R.

### The most representative predictive model

The analyses described above was repeated 1000 times (in the case of fHLR and bHLR, 5000 times since it is a less computationally heavy analysis) with different data split by varying the seed randomly to control the potential error induced by the split. A mode from the R-squared distribution, derived from the most representative model, was used to describe model fitness (see Figure 2 in the supplementary materials for the analysis flow). The top 3 most important Disconnectome components (https://identifiers.org/neurovault.collection:7735) were displayed using Surfice (https://www.nitrc.org/projects/surfice/). See Table 2 from the Supplementary material for the component names and thresholds used to display them.”

**Figure 2.**
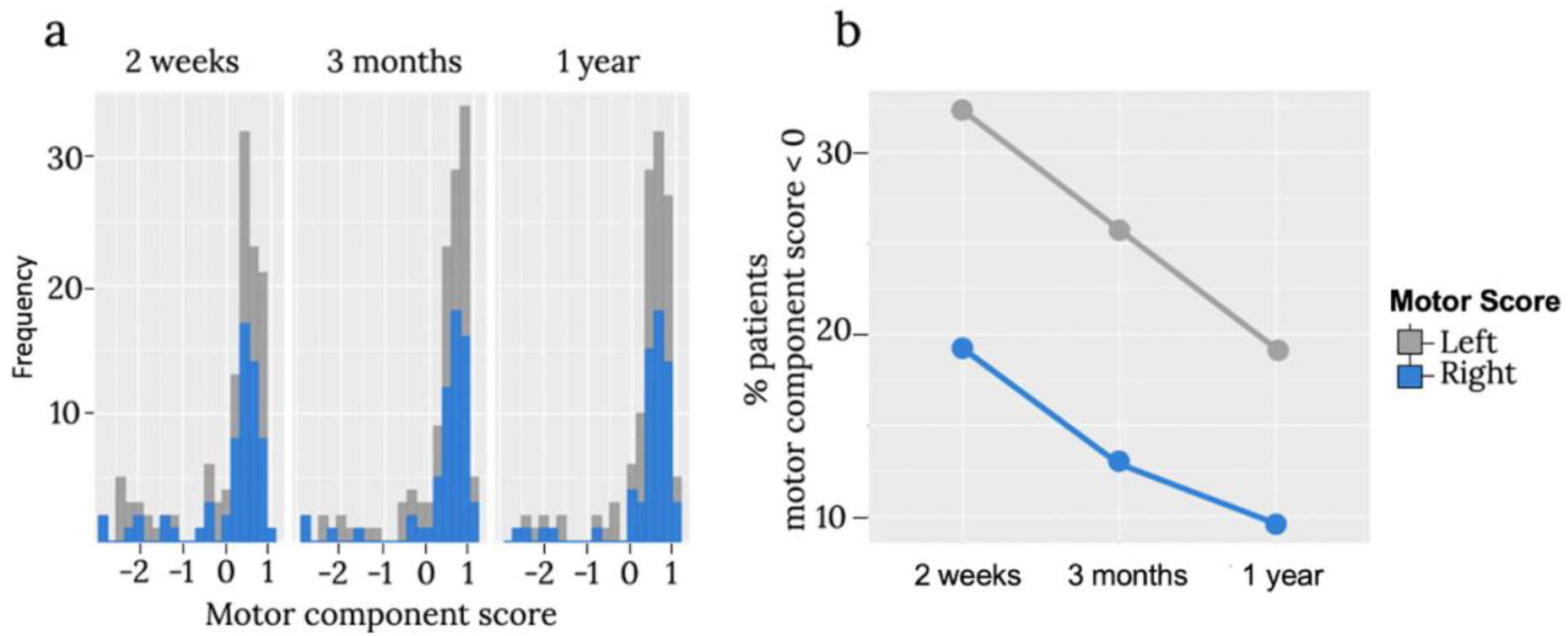
The distribution of motor scores across two weeks, three months, and one-year periods after stroke. a) Overall distribution of the score, b) Proportion of patients with a motor score <0.

### Anatomy

The white matter was identified and labelled manually by expert anatomists (MTS/SJF) according to the Atlas of Human Brain Connections (Rojkova et al. 2016; Thiebaut de Schotten et al. 2015).

## Results

### Lesion characteristics (location, overlay)

Patients’ lesions and disconnection maps from the dataset (N = 62) were distributed bilaterally with 67% of lesions in the right hemisphere (Figure 1). The most common site of damage was observed in the right subcortical regions (n=27; other areas n<15), including the thalamus, putamen, caudate, pallidum, hippocampus, amygdala, nucleus accumbens, insula, subcallosal cingulate, paracingulate, and parahippocampal areas. The disconnection maps generated with the individual lesions mirrors this bilateral but right prevalent pattern of disconnection with the highest overlap involving the ventral visual pathways, internal capsule, and perisylvian white matter (Catani and Thiebaut de Schotten 2012).

### Behavioural characteristics (Motor impairment)

Figure 2 indicates that a significant portion of patients in our sample presented with motor impairments (motor score <0). The proportion of patients with motor disabilities reduced from two week to one year after the stroke in line with previous findings (Hatem et al. 2016).

### Prediction of the motor outcomes

Linear regressions used the patients’ estimated scores of each component to predict motor scores’ components (the left and right side of the body, n = 2) two weeks, three months, and one year after the stroke (n = 3 time points). We performed different linear algorithms (backward/forward hierarchical regressions, backward/forward stepwise regression, lasso and ridge regressions) to select the most important components that are able to explain as much variance in new data as possible. Each regression was repeated 1000 times (5000 times in case of the hierarchical regressions) with different training and testing dataset splits.

In comparison to other algorithms, the predictions of the ridge regression are not superior at any time point (2 weeks, 3 months, 1 year) for the left side motor impairments. However, it demonstrated robust prediction power for the right side motor impairments (Figure 3, 4).

**Figure 3.**
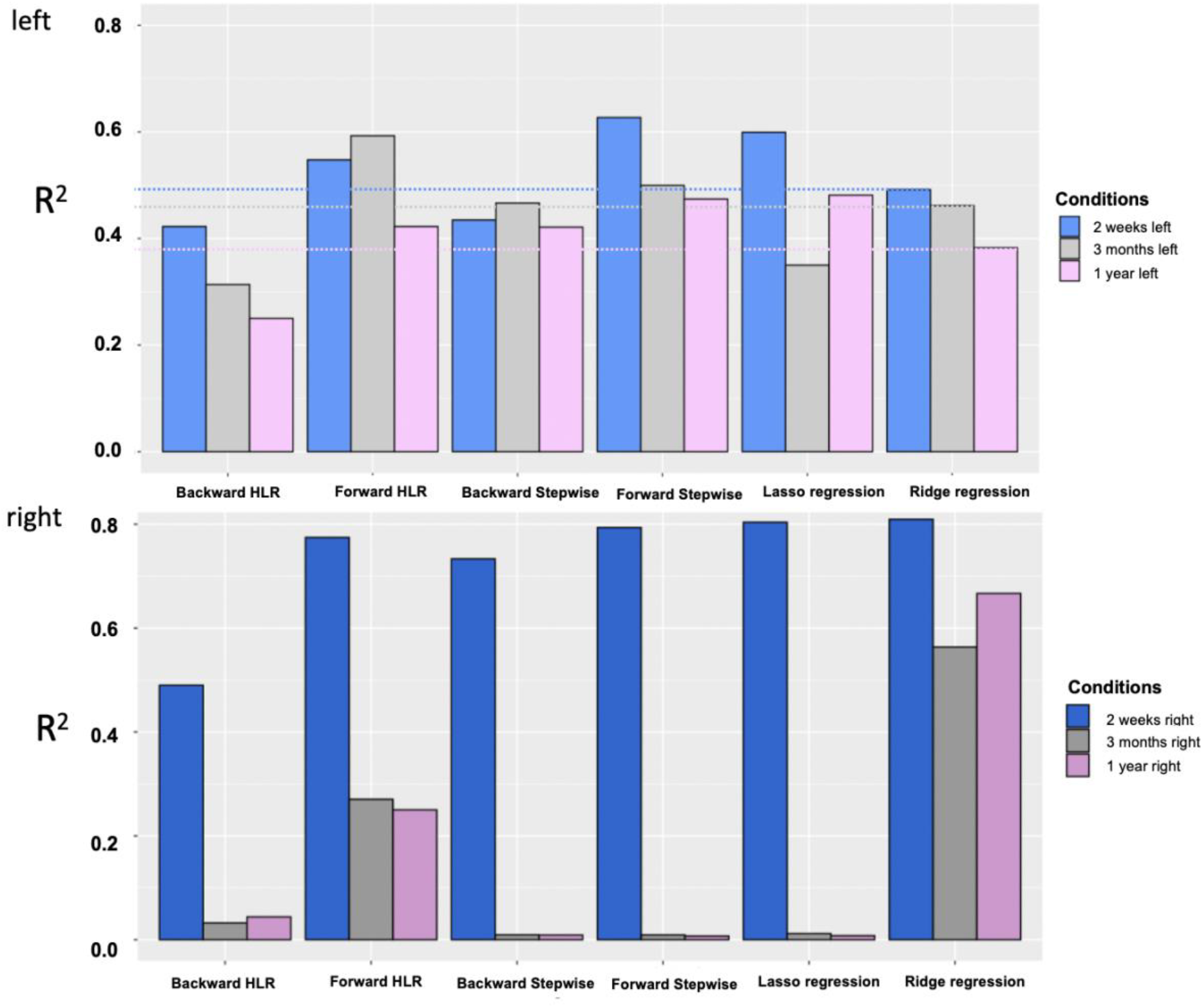
The comparison of different model predictions (R2) on unseen data.

**Figure 4.**
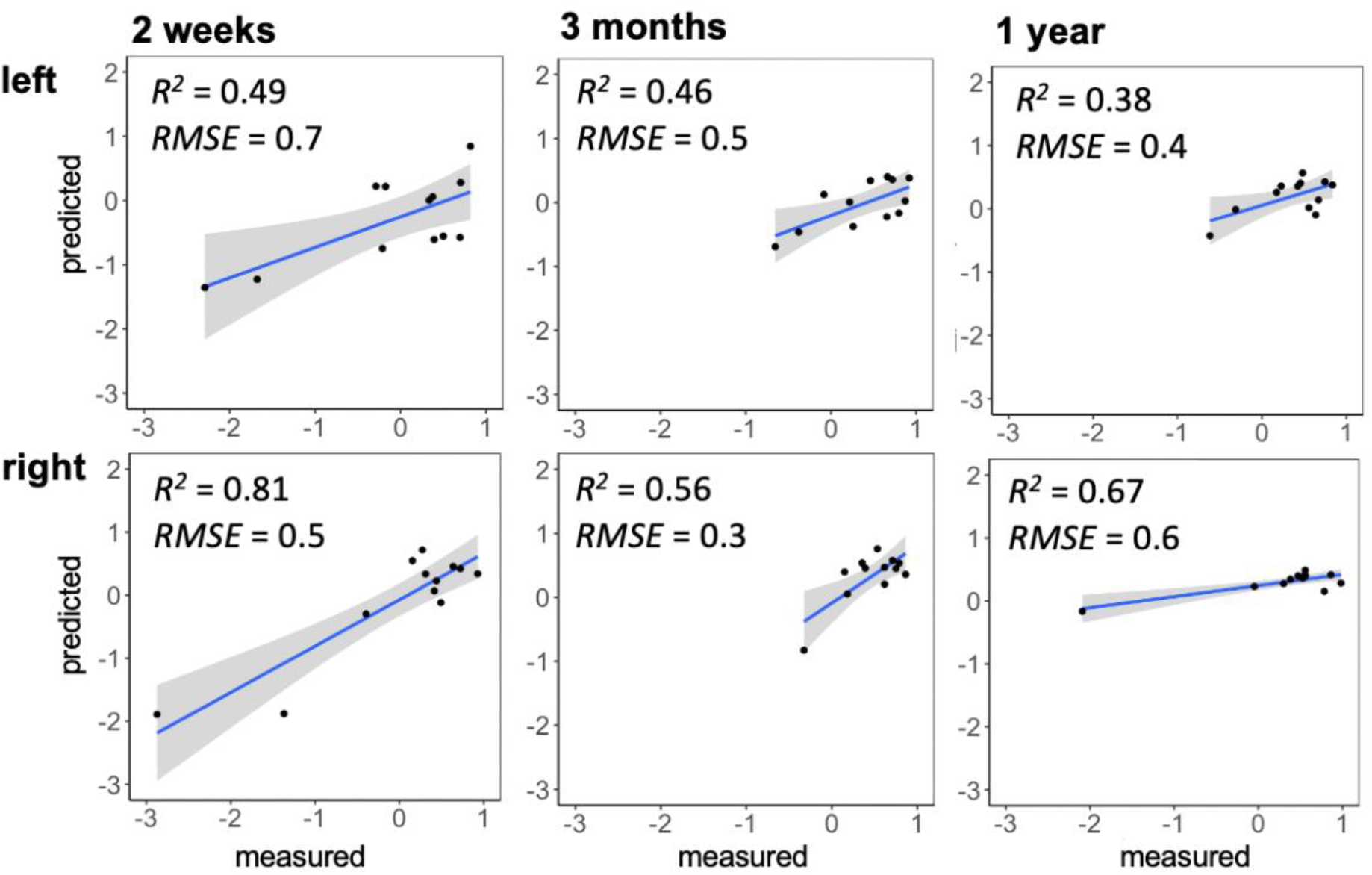
Prediction accuracy (R2) of the ridge regression models in the testing set (20% of the data, 12 patients), based on the mode of R distribution (1000). Left and right indicate motor scores (see Figure 3 from the Supplementary materials for the predictions of other approaches implemented in the study).

The ridge regression does not eliminate predictors from the model. However, some component coefficients that are considered to be less important become close to 0 due to the L2-regularisation penalty. Here we demonstrated the top 3 most important components in predicting individual motor scores at 3-time points (Figure 5).

**Figure 5.**
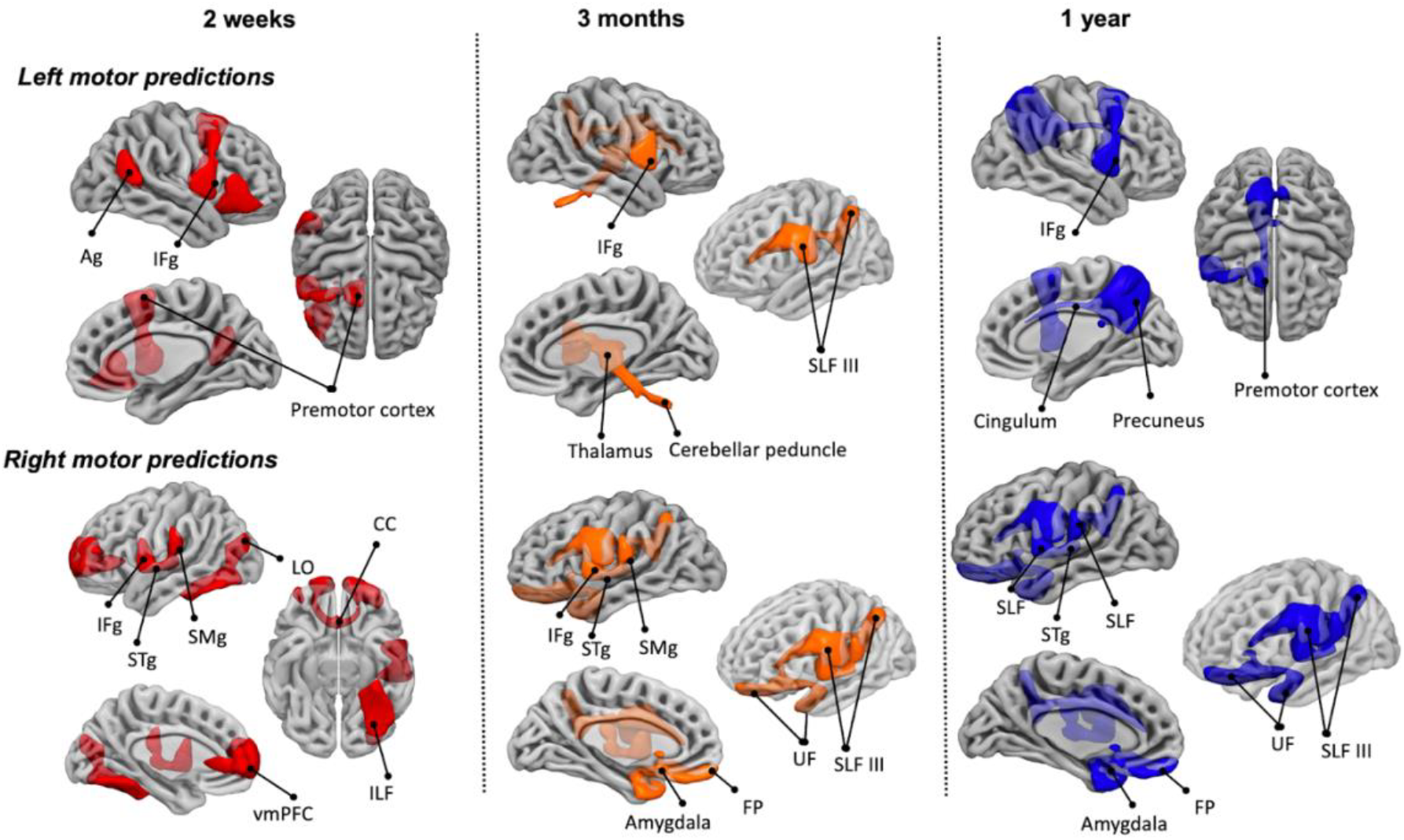
Top-3 contributing components in ridge regression at three time points after stroke onset (two weeks, three months, and one year) for motor impairments. SLF: Superior longitudinal fasciculus, IFg: Inferior frontal gyrus, SMg: supramarginal gyrus white matter, Ag: Angular gyrus, STg: Superior temporal gyrus, vmPFC: ventro-medial prefrontal cortex, ILF: Inferior longitudinal fasciculus, SMg: supramarginal gyrus, FP: frontal pole, UF: Uncinate fasciculus, LO: Lateral occipital gyrus, CC: corpus callosum.

The disconnection of the component corresponding to the right frontal gyrus was a significant contributor in explaining the variance for the left motor impairments starting from 2 weeks to 1 year after stroke. The involvement of the premotor cortex was prominent at 2 weeks and 1 year. At 3 months it was surpassed by more subcortical regions such as the thalamus, and cerebellar peduncle, although it was still chosen to be in the top-10 contributing components by the ridge regression model (see Figure 5 from the Supplementary materials for the predictor importance plots). Surprisingly, the left superior longitudinal fasciculus III were in the top 3 components that predict left motor impairment at 3 months.

Correspondingly, right motor predictions at 3-time points were mainly driven by the disconnection of left inferior frontal gyrus together with the superior temporal gyrus, and the supramarginal gyrus disconnection. The orbitofrontal regions disconnection together the superior longitudinal fasciculus III with and the uncinate started to predict individual motor scores significantly 3 months after the stroke and were replicated 1 year after stroke.

## Discussion

In this study, we predicted motor impairments across different time points based on the pattern of acute brain disconnections two weeks after a stroke. Our analysis reveals three primary results. First, brain disconnection patterns can accurately predict motor impairment. Second, the disconnection patterns leading to impairment are not the same at the two weeks, three months, and one year after a stroke. Third, the predictions were replicable, to some extent, in the cross-validation analysis. Overall, the results indicate that while some plasticity mechanisms exist changing the structure-function relationship, early disconnection patterns prevail when predicting motor impairment at different times after the stroke.

Prediction of behavioural impairments based on brain data has been one of the early goals of clinical neuropsychology. Pioneer scientists analysed neuropsychological impairments of single cases and tried to link them with brain lesions (e.g. patient Leborgne with a lesion in the left inferior frontal gyrus and loss of articulation). While this approach has offered fruitful discussions on the link between a specific region in the brain and a cognitive function (i.e. localisationism), it is hard to use this approach to accurately predict individual patients in the clinic because of variations in lesion characteristics and individual brain variations. These structural and functional differences in healthy brains add extra variability and, therefore, are an additional challenge to generalise a single case to the whole patient population (Smith et al. 2019). Another extremity of analysing lesion-symptom relationships is the group-level analysis, where all data is averaged to a common space. This diminishes the role of interindividual variability (Forkel et al. 2021), and inference about an individual based on group-level data analysis leads to a potential error, known in the literature as “ecological fallacy” (Portnov et al. 2007; Robinson 1950). Therefore, predictions based on these two approaches could be misleading. With the availability of big datasets and the development of new statistical tools, it became possible to advance from single cases and group-level studies toward an integrative approach that accounts for interindividual variations but generalises to the group-level. This integration allows researchers to make personalised and individual predictions like in the present study.

Preliminary attempts used different machine learning approaches to predict motor impairment. However, most of the studies fall into one of the following pitfalls. The models are not validated on an independent dataset. They also focus on the lesion location, statistically ignoring the remote effects of a lesion on distant brain regions. Finally, they only consider a few aspects of motor functions independently rather than the entire pattern of motor abilities. Akin to Salvalaggio et al. (2020), we considered these issues that may downplay the potential power of prediction motor symptoms based on neuroimaging data. We applied the resampling method to validate the results. We used behavioural data that includes excellent variability of neuropsychological tests that capture motor disorders from different angles. We applied disconnection analysis that is a more accurate statistical measure of the theoretical concept of diaschisis. However, in contrast to Salvalaggio and colleagues’ (2020) work, we used the Disconnectome (Thiebaut de Schotten et al. 2020) developed on an independent and much larger cohort of stroke patients dataset (n=1333) to model the profile of disconnection of individual patients. The Disconnectome allowed increasing the predictive power for motor impairments from 37% to 49% (right hemisphere lesion) and from 42% to 81% (left hemisphere lesion) at 2 weeks. These progresses are indicative of the importance of the disconnectome in the prediction of neuropsychological impairment and a significant step forward personalised prognosis in clinical neuropsychology. Additionally, we compared the results of our approach with previously established method such as lesion load analysis of cortico-spinal tract and our algorithm proved to explain more variance (see Figure 7 from the Supplementary materials).

In addition, it is worth noting that in the current study, we predict long-term symptoms at one year. A long-term prediction at one year based on acute brain imaging is challenging as additional factors may interact with the recovery (e.g., intensive rehabilitation, plasticity), which will have to be studied further before this work can be routinely incorporated into a clinical routine. For instance, in the current study, we observed a drop in the model’s ability to capture the variance in right-sided motor scores of patients from 81% (2 weeks) to 67% (1 year). This might be due to the variability of brain plasticity capacities and functional coping strategies across patients. Additional factors such as, for example, patients’ demographics (e.g. age, education) might increase the model’s accuracy for long-term symptom predictions in future research.

Studying motor impairments longitudinally allowed us to indirectly assess plasticity mechanisms over time. We demonstrated different patterns of disconnections responsible for motor impairments at two weeks, three months, and one year after a stroke. Particularly some components that initially were less responsible for the motor impairment (two weeks after the stroke) play greater role later in recovery. For instance, left sided motor deficits become well predicted by cingulum and precuneus after 1 year. Those regions were previously linked with a modulatory effect on motor areas i.e., on primary motor cortex (Wenderoth, Debaere, Sunaert & Swinnen, 2005). In addition, our result is consistent with the finding of Takenobu and collegaures (2014) where the fractional anisotropy value in cingulum was positively correlated with motor function recovery after stroke (Takenobu et al, 2014). Another research group demonstrated that contralateral cingulate cortex and cerebellar activation is associated with improved motor function which is in line with the results of our study as well (Tong et al, 2016). It seems that the recruitment of regions beyond the motor cortex could be one of the plasticity mechanisms of recovery from stroke that damages primarily a part of cortico-spinal tract. The inferior frontal gyrus could be another example. It was one of the main contributors in prediction of motor impairment both for the left and right sided lesions. Tang et al (2015) demonstrated that the focal motor pathway stroke extends to regions beyond traditional motor-related networks such as the inferior frontal gyrus since they found a decreased inter-hemispheric connectivity in the inferior frontal gyrus and middle temporal gyrus in patients after stroke (>3 months) in comparison to healthy controls (Tang et al, 2015).

Although the current study allowed us to predict motor outcomes of patients at different time points after a stroke, the model based on ridge regression still requires improvements, when predicting long-term symptoms. We believe that adding other factors (e.g. demographic, clinical, socioeconomic variables) that likely interact with the recovery of patients and using larger sample size can help us increase the model’s predictive power. For example, there is evidence that a multimodal approach can outperform single-modality-based algorithms in the discrimination of patients (Lu et al. 2018). In this instance, different modalities complement and confirm each other and thus, this redundancy coming from various sources allows the algorithm to better estimate the prediction. Adding age, sex, lesion volume, or resting-state functional MRI data as an additional modality, for example, could provide valuable information to the algorithm’s prediction accuracy during training. Despite these advantages, multimodal prediction algorithms are rarely clinically feasible and often too expensive for large-scale longitudinal studies in terms of acquisition, computing powers, and analysis time. Further the purpose of this investigation was to assess the sole contribution of brain disconnection to the prediction of motor performance longitudinally after a stroke, not the contribution of other factors to build the model that best fits (but see Figure 6 from the Supplementary materials where we included age and total lesion load as additional predictors).

We applied different linear methods (i.e. stepwise, hierarchical, Lasso, Ridge) to predict long-term motor scores in patients (see Figure 4 from the Supplementary materials) and we observed that retained components varied across different models. For example, comparing stepwise and hierarchical regressions, both backwards and forwards, demonstrated poor consistency: only few (less than 3 usually) components were retained in the final models between the different approaches (see Table 3 from the Supplementary materials). The possible reason is the existence of correlated components. Stepwise and hierarchical regression models do not account for the multicollinearity problem that exists in our data in comparison to ridge and lasso regressions that use regularisation techniques. In the case of the Lasso regression, only one of the highly collinear predictors usually stay in the model. This solution to the multicollinearity problem is not suitable for our task and the final model might be misleading in interpretation. For example, if a component that includes motor regions is highly correlated with a component that includes cortico-spinal-tract, then only one of them will be retained in the final lasso regression model. Nevertheless, we need to know that both of them play an important role in predicting motor impairments and by dropping one of them will make our interpretation of the brain region involvements incomplete., By contrast, ridge regression appeared to be the most reliable and suitable method to predict motor outcomes in our dataset since (1) it accounts for multicollinearity, (2) it did accurately predict longitudinal right motor impairments better than other methods, and (3) it demonstrated the consistency of the important components with Lasso regression and across time (2 weeks, 3 months, 1 year). For example, out of the top 20 important components in the ridge regression model, 16 components were coinciding with the components retained in the lasso regression for the right motor impairment at 2 weeks (see Figure 5 from the Supplementary materials). Another possible reason for the inconsistency in the retained components is the small sample size. This has been one of the major problems in the application of machine learning algorithms on small datasets. However, the nested cross-validation technique must have diminished the biased effect and led to a higher consistency across the results (Vabalas et al, 2019).

Another potential limitation of the study is that the disconnection maps of 6 haemorrhagic stroke patients (∼10% of the dataset) were described with the Disconnectome that was developed only on ischemic stroke patients. However, the Disconnectome was able to explain the variance in those 6 patients’ disconnection maps as well (R2: M = 0.983, SD = 0.011). Thus, the inclusion of 6 patients with haemorrhagic strokes did not influence our results.

Overall, we managed to increase the model’s prediction quality two weeks after a stroke using the Disconnectome developed in an independent cohort of patients. This is a big step toward creating a clinical tool that will be able to complement prognosis of motor symptoms recovery in individual patients. We believe that using a multimodal approach in future studies (e.g. the inclusion of additional factors) and increasing the cohort size will allow the model to make more accurate long-term personalised predictions which, in turn, will inform tailored rehabilitation pathways. This achievement would only be possible within an open science framework capitalising on an effort to form collaborations between neurological centres to pool data.

## Data Availability

The code used to analyse the data is openly available from (GitHub link)

https://github.com/lidulyan/Hierarchical-Linear-Regression-R-

## Acknowledgments

We thank the University of Bordeaux and CNRS for the infrastructural support. This project has received funding from the European Research Council (ERC) under the European Union’s Horizon 2020 research and innovation programme (grant agreement No. 818521), the Marie Skłodowska-Curie programme (grant agreement No. 101028551), The Erasmus+ for Traineeships Programme, and the Donders Mohrmann Fellowship No. 2401515 (SJF, NEUROVARIABILITY).

## Data availability

The forward/backward hierarchical linear regression, backward/forward stepwise regression, lasso and ridge regressions was carried out in RStudio (v.1.3.1093; RStudio Team, 2020). The code is freely available (https://github.com/lidulyan/Stepwise-Lasso-Ridge and https://github.com/lidulyan/Hierarchical-Linear-Regression-R-). The code for the estimation of the component’s score is freely available (https://github.com/lidulyan/Estimate_CompScore_lm).

## Notes

### Competing Interest Statement

The authors have declared no competing interest.

### Funding Statement

This project has received funding from the European Research Council (ERC) under the European Union Horizon 2020 research and innovation programme (grant agreement No. 818521), the Marie Sklodowska-Curie programme (grant agreement No. 101028551), The Erasmus+ for Traineeships Programme, and the Donders Mohrmann Fellowship No. 2401515 (SJF, NEUROVARIABILITY).

### Author Declarations

Ethics committee/IRB of the Washington University gave ethical approval for this work

### Summary of Updates

We amended the manuscript according to the reviewers feedback.

