## Supplementary material for "Longitudinal prediction of motor dysfunction after stroke: a disconnectome study"

### Glossary

*The Disconnectome* includes 46 components that have been parcellated into 372 anatomical regions (it is a matrix of 46 x 372) and derived from a dataset of 1333 patients.

A *disconnection pattern/map* is a map/pattern of white matter pathway disconnection probabilities and its impact on other brain areas of a patient (i.e. it is a matrix of 1 x 372 regions).

"*Disconnectome Maps*" is the name of the function in the BCBToolkit (toolkit.bcblab.com) that computes a disconnection pattern/map for a patient

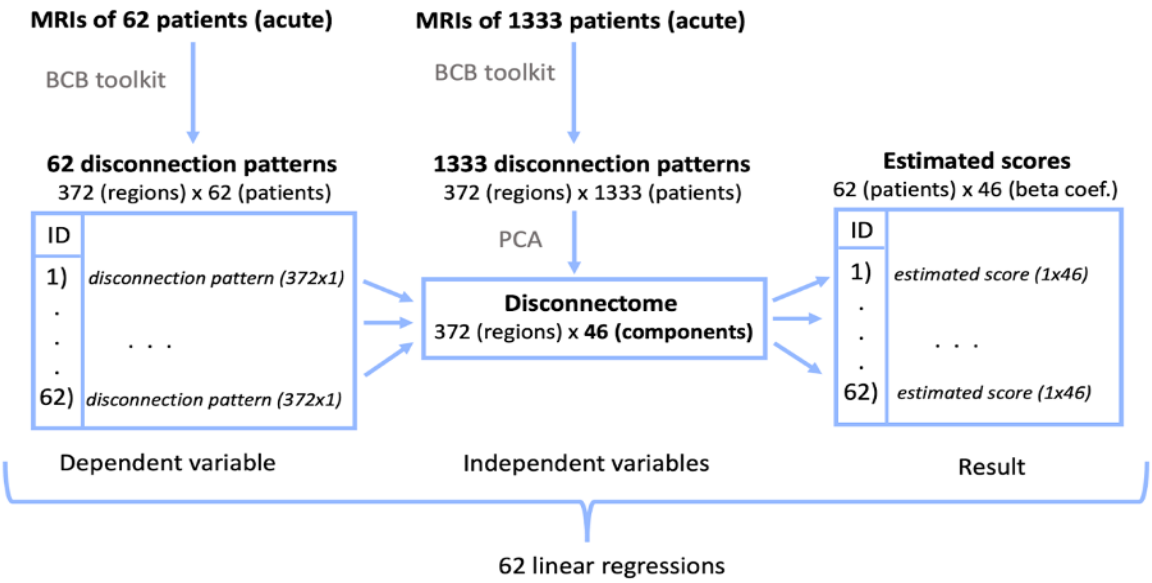


**Figure 1.** 62 linear regressions were performed to describe the disconnection patterns of 62 patients with the Disconnectome.

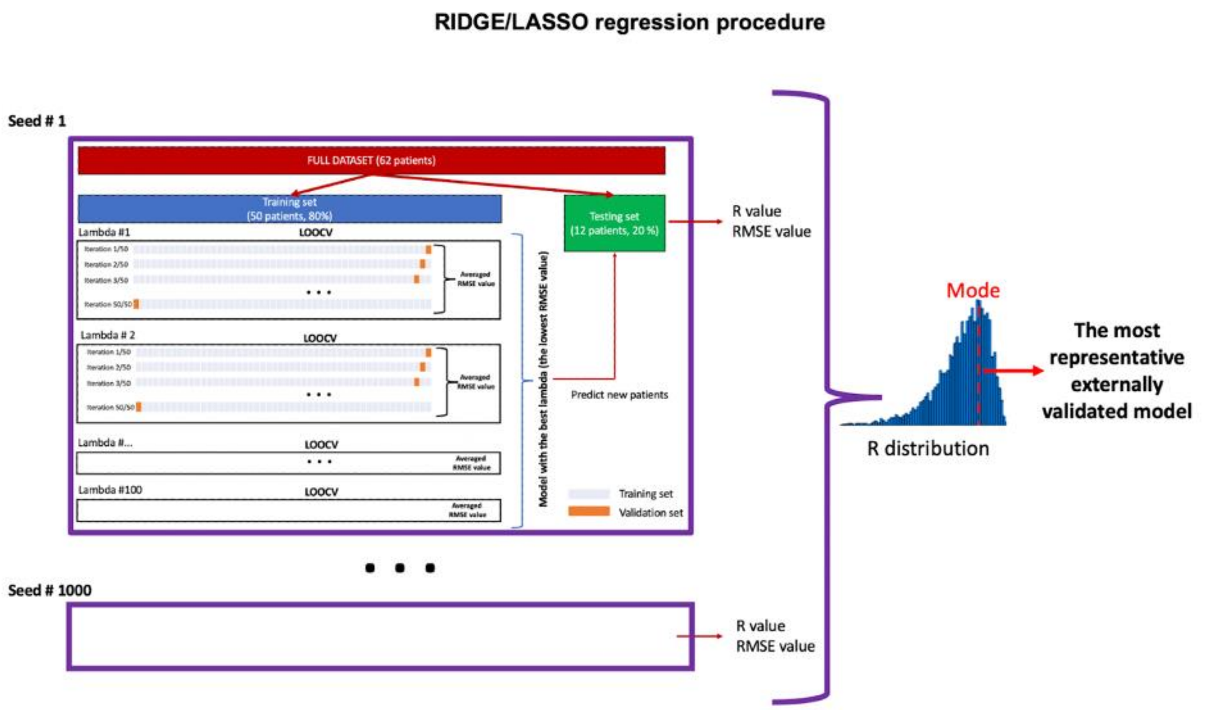


**Figure 2.** The flow of analysis for the ridge and lasso regressions. RMSE: root mean square error, LOOCV: leave-one-out cross-validation, Lambda – L1/L2 regularization value.

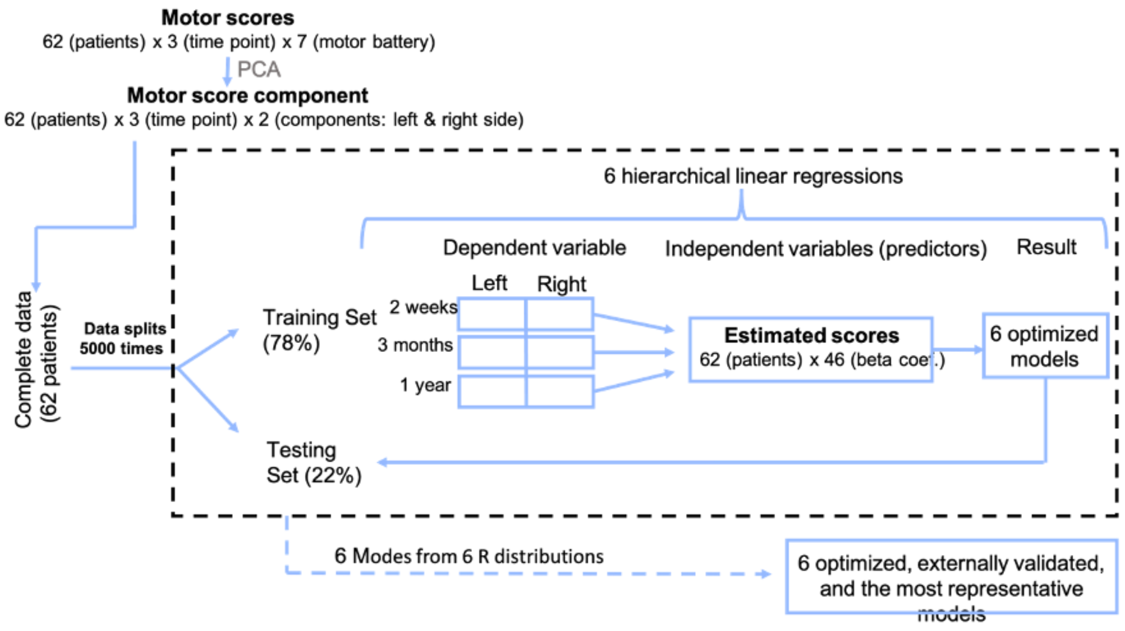


**Figure 3.** The flow of analysis for hierarchical linear regressions.

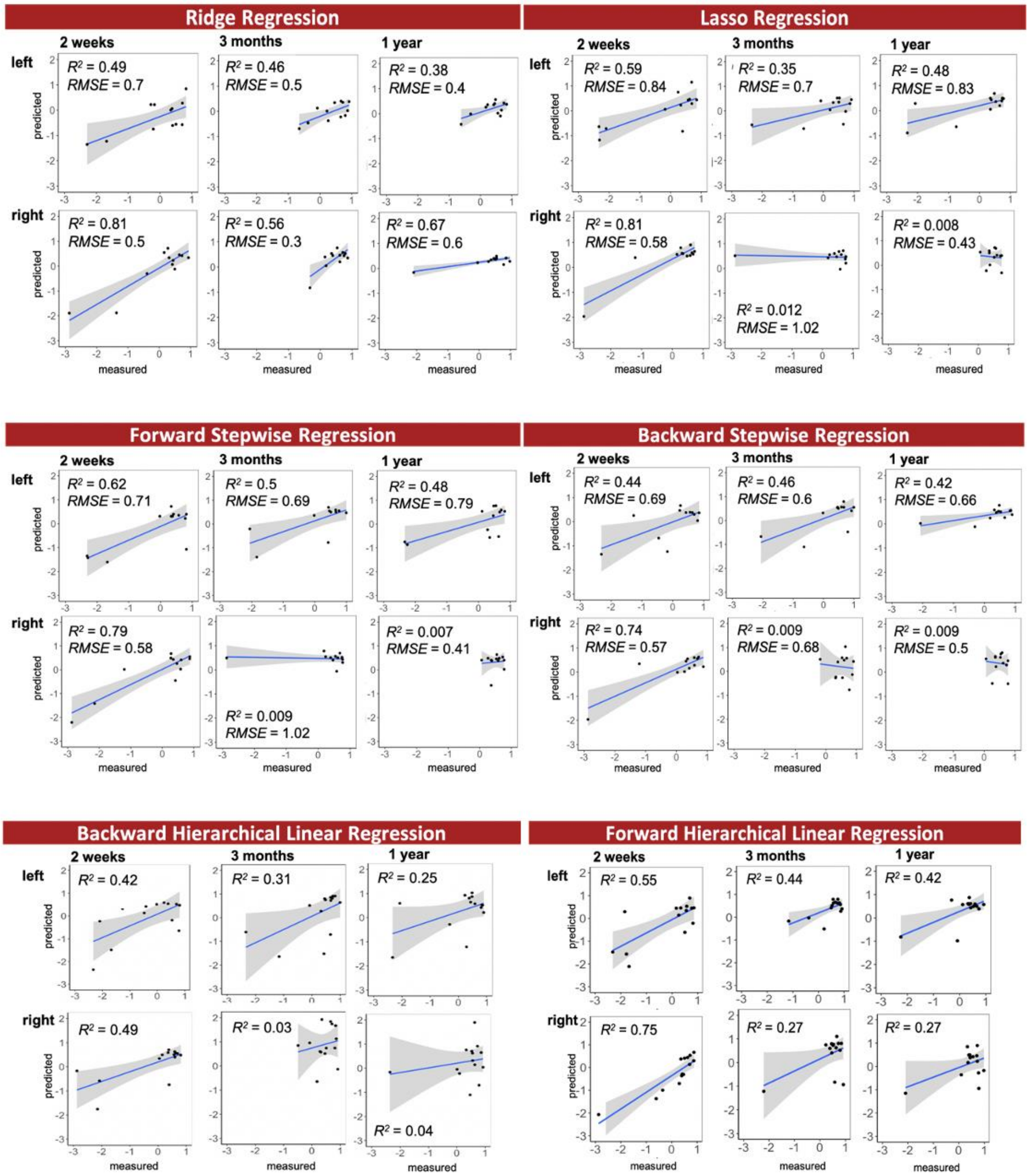


**Figure 4.** Prediction accuracy ($R^2$) of different algorithms in the testing set, based on the mode of R distribution. Left and right indicate motor scores.

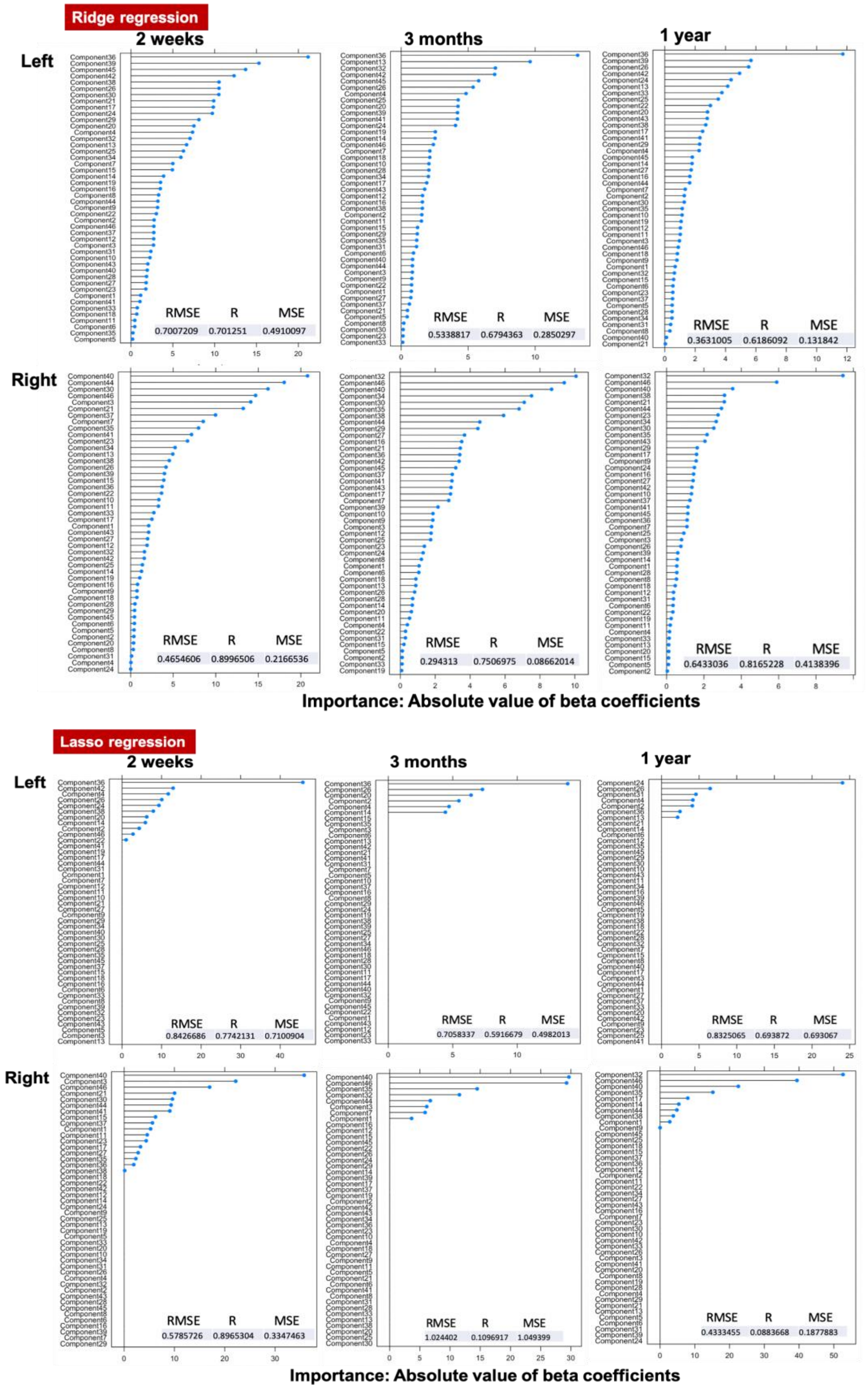


**Figure 5.** Predictor importance plots for ridge and lasso regressions per each condition

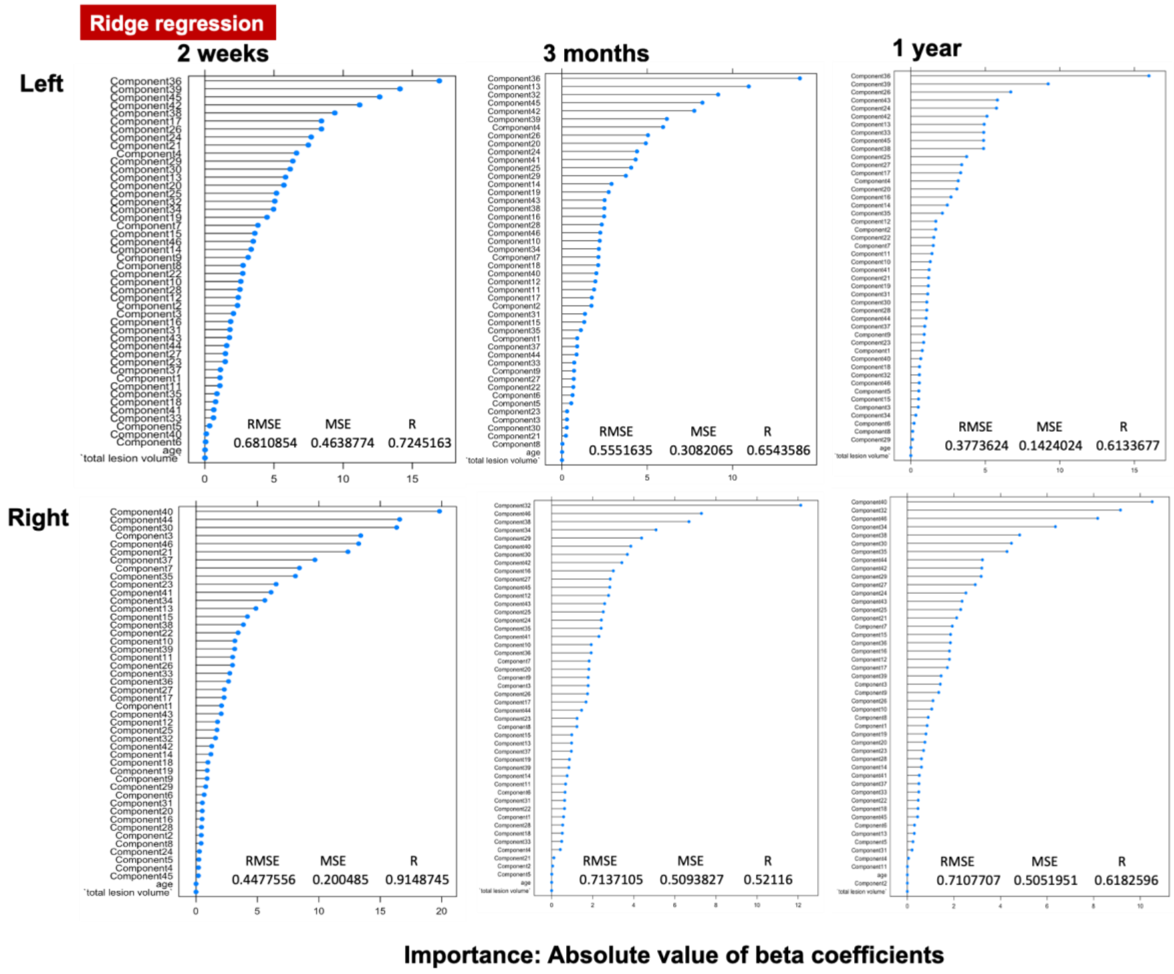


**Figure 6.** Predictor importance plots for ridge regressions per each condition. Age of the patients and total lesion volume were included in regressions and demonstrated low importance in explaining the variance in data.

We compared our method to a previously established method to estimate the corticospinal tract lesion load (e.g. tractotron, www.bcbtoolkit.com). Using this method we predict the motor outcomes using a probability of cortico spinal tract (CST) disconnection (Figure 7). The probability was calculated with a tractotron as implemented in the BCBtoolkit (toolkit.bcblab.com) . The results showed *that the* lesion load to the left CST was a reliable predictor for right motor impairments and the right CST for the left motor impairments. In comparison, however, the algorithm employed in our study explains more *variance* in the observed motor behaviour across all three time points (2 weeks, 3 Months, 1 year). This applies to the right and the left motor impairments.

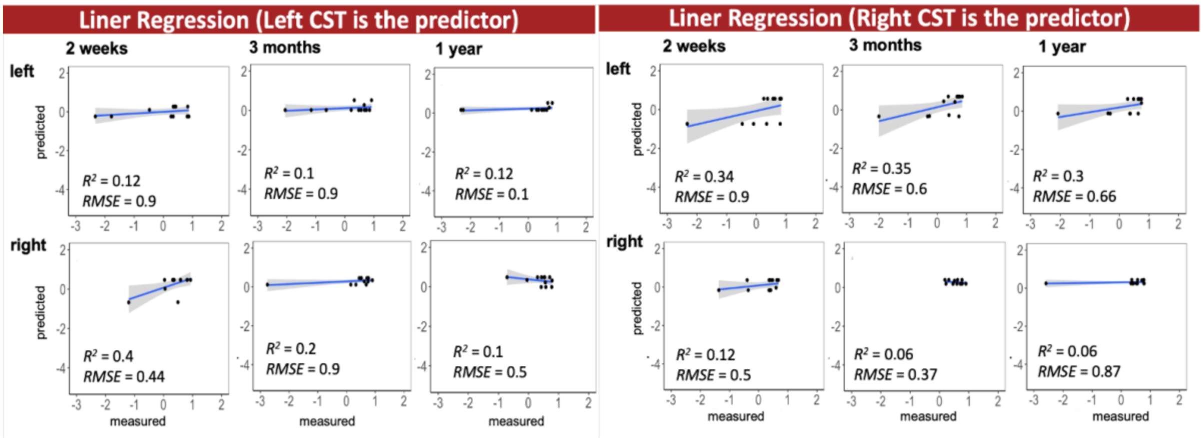


**Figure 7.** Prediction accuracy ($R^2$) of the linear regression models in the testing set (20% of the data, 12 patients), based on the mode of R distribution (1000). Left and right indicate motor scores. Independent variable: probability of cortico-spinal tract (CST) disconnection

**Example: Patient FSC_040**

Multiple linear regression analysis result: R^2 adj = .97, F(46, 325) = 270.5, p < .001

**Table 1.** Power calculations of the multiple linear regression analysis of the patient FSC_040.

**F tests** Linear multiple regression: Fixed model, $R^2$ deviation from zero

**Analysis** Post hoc: Compute achieved power

**Input**

| | |
|---|---|
| Effect size $f^2$ | 33,36426 |
| α err prob | 0,05 |
| Total sample size | 372 |
| Number of predictors | 46 |

**Output**

| | |
|---|---|
| Noncentrality parameter λ | 12411,5 |
| Critical F | 1,4049935 |
| Numerator df | 46 |
| Denominator df | 325 |
| Power (1-β err prob) | 1 |

**Table 2.** The 3 top the most important Disconnectome components https://identifiers.org/neurovault.collection:7735 displayed with Surfice https://www.nitrc.org/projects/surfice/ and used thresholds for each condition.

| Condition | Component | Threshold |
|---|---|---|
| Acute left | componentA 45 | neg8 |
| | componentA 39 | 8 |
| | componentA 36 | 15 |
| acute right | componentA 44 | 10 |
| | componentA 40 | 10 |
| | componentA 30 | 11 |
| 3months_left | componentA 36 | 15 |
| | componentB 13 | 10 |
| | componentA 32 | neg5 |
| 3months_right | componentA 46 | neg15 |
| | componentA 40 | 10 |
| | componentA 32 | neg5 |
| 1year_left | componentA 26 | 12 |
| | componentA 39 | 8 |
| | componentA 36 | 15 |
| 1year_right | componentA 46 | neg15 |

| | | |
|---|---|---|
| | componentA 40 | 10 |
| | componentA 32 | neg5 |

**Table 3.** Retained components for the forward/backwards hierarchical linear regressions, forward/backward stepwise regressions

| Cond. | Regression type | Type of validation | # | Retained components |
|---|---|---|---|---|
| 2weeks left | Backward HLR | 5000 permutations with testing on 22% data | 9 | Component40, Component38, Component37, Component31, Component29, Component21, Component18, Component14, Component2 |
| 2weeks right | Backward HLR | 5000 permutations with testing on 22% data | 4 | Component42, Component39, Component15, Component3 |
| 3m left | Backward HLR | 5000 permutations with testing on 22% data | 7 | Component37, Component35, Component26, Component21, Component14, Component4, Component2 |

| | | | | |
|---|---|---|---|---|
| 3m right | Backward HLR | 5000 permutations with testing on 22% data | 25 | Component42, Component40, Component35, Component33, Component32, Component30, Component29, Component28, Component26, Component25, Component22, Component21, Component20, Component17, Component16, Component15, Component14, Component13, Component11, Component10, Component8, Component7, Component5, Component3, Component2 |
| 1y left | Backward HLR | 5000 permutations with testing on 22% data | 8 | Component28, Component26, Component24, Component18, Component16, Component13, Component7, Component2 |
| 1y right | Backward HLR | 5000 permutations with testing on 22% data | 7 | Component32, Component30, Component27, Component24, Component19, |

| | | | | |
|---|---|---|---|---|
| | | | | Component17, Component12 |
| 2weeks left | Forward HLR | 5000 permutations with testing on 22% data | 3 | Component2, Component8, Component24 |
| 2weeks right | Forward HLR | 5000 permutations with testing on 22% data | 5 | Component1, Component3, Component15, Component23, Component40 |
| 3m left | Forward HLR | 5000 permutations with testing on 22% data | 3 | Component21, Component14, Component36 |
| 3m right | Forward HLR | 5000 permutations with testing on 22% data | 4 | Component1, Component7, Component14, Component35 |
| 1y left | Forward HLR | 5000 permutations with testing on 22% data | 4 | Component2, Component9, Component24, Component36 |
| 1y right | Forward HLR | 5000 permutations with testing on 22% data | 3 | Component9, Component32, Component46 |

| | | | | |
|---|---|---|---|---|
| 2weeks left | Backward Stepwise | LOOCV on training set (80%) and test on 20% with 1000 permutations | 1 | Component2 |
| 2weeks right | Backward Stepwise | LOOCV on training set (80%) and test on 20% with 1000 permutations | 5 | Component1 Component3 Component5 Component37 Component44 |

| | | | | |
|---|---|---|---|---|
| 3m left | Backward Stepwise | LOOCV on training set (80%) and test on 20% with 1000 permutations | 2 | Component2 Component25 |
| 3m right | Backward Stepwise | LOOCV on training set (80%) and test on 20% with 1000 permutations | 5 | Component18 Component27 Component29 Component35 Component44 |
| 1y left | Backward Stepwise | LOOCV on training set (80%) and test on 20% with 1000 permutations | 2 | Component2 Component24 |
| 1y right | Backward Stepwise | LOOCV on training set (80%) and test on 20% with 1000 permutations | 3 | Component17 Component32 Component46 |
| 2weeks left | Forward Stepwise | LOOCV on training set (80%) and test on 20% with 1000 permutations | 1 | Component2 |
| 2weeks right | Forward Stepwise | LOOCV on training set (80%) and test on 20% with 1000 permutations | 4 | Component3 Component7 Component21 Component40 |
| 3m left | Forward Stepwise | LOOCV on training set (80%) and test on 20% with 1000 permutations | 2 | Component2 Component24 |
| 3m right | Forward Stepwise | LOOCV on training set (80%) and test on 20% with 1000 permutations | 4 | Component1 Component35 Component40 Component46 |
| 1y left | Forward Stepwise | LOOCV on training set (80%) and test on 20% with 1000 permutations | 1 | Component2 |
